# SDoH-Aware Approach to Prostate Cancer Screening: Addressing Overdiagnosis of Prostate Cancer using PSA

**DOI:** 10.1101/2024.07.31.24311297

**Authors:** Ashley Lewis, Yash Samir Khandwala, Tina Hernandez-Boussard, James D Brooks

**Author notes:** Shared senior authorship (Tina Hernandez-Boussard, James D Brooks).

## Abstract

This study investigates the potential of multimodal data for prostate cancer (PCa) risk prediction using the All of Us (AoU) research program dataset. By integrating polygenic risk scores (PRSs) with diverse clinical, survey, and genomic data, we developed a model that identifies established PCa risk factors, such as age and family history, and a novel factor: recent healthcare visits are linked to reduced risk. The model’s performance, notably the false positive rate, is improved compared to traditional methods, despite the lack of Prostate-Specific Antigen (PSA) data. The findings demonstrate that incorporating comprehensive multimodal data from AoU can enhance PCa risk prediction and provide a robust framework for future clinical applications.

**Code Available:** https://github.com/ashlew23/pc_multimodal

## 1. Introduction

Prostate cancer (PCa) is one of the most common malignancies affecting men worldwide and it is a complex and multifaceted disease characterized by significant heterogeneity and disparities among individuals.^1^ While prostate-specific antigen (PSA) testing has undoubtedly played a crucial role in early detection, it has also been associated with significant overdiagnosis and overtreatment, leading to unnecessary biopsies, psychological stress, and potential treatment-related complications.^2^ To address this, novel methods that use a precision-medicine approach to improve traditional PSA testing are needed to identify cases that might be missed by conventional biological markers.^3^ Integrating genetic information with clinical and lifestyle data presents a promising avenue for advancing disease prediction and precision medicine.

Polygenic risk scores (PRSs) quantify genetic predisposition by analyzing multiple genetic variants and have emerged as a promising tool for PCa screening.^4^ These scores offer a promising avenue for identifying men at higher genetic risk of developing PCa, thereby refining screening strategies and potentially reducing overdiagnosis. However, their use can sometimes lead to overdiagnosis, and they may not perform as well as validated PCa biomarkers and therefore the utility of PRSs in clinical practice remains uncertain, particularly when compared to traditional factors such as Social Determinants of Health (SDoH).^5^ SDoH, encompassing socioeconomic status, education, access to healthcare, and geographic location, significantly influence health outcomes and access to medical interventions.^6^

This study aims to evaluate the contributions of PRSs with SDoH, to develop an integrated, multimodal approach that leverages both genetic and social factors to optimize screening strategies. This SDoH-aware approach not only aims to improve screening issues associated with PSA but also strives to enhance health equity by ensuring that screening practices are informed by the broader social context in which patients live. The All of Us (AoU) research program offers a new opportunity to evaluate this hypothesis, providing access to both multimodal data and a large, diverse research population. This framework can be applied to other disease types to inform SDoH-aware screening practices. Understanding the relative importance of each data type in predicting disease incidence can help refine predictive models and identify key risk factors.

## 2. Related Work

### 2.1. Modern Methods for Prostate Cancer Incident Prediction

PSA-only based screening has significantly contributed to the overdiagnosis of PCa, leading to controversy regarding its utility as a biomarker. In 2018, the US Preventive Services Task Force highlighted that periodic PSA-based screening might lead to false positives, which could require additional testing and potential biopsies, thereby posing risks of overdiagnosis and unnecessary surgical treatment.^7^ Beyond the immediate physical implications, overdiagnosis has been associated with both short-term and long-term anxiety.^8^ These concerns underscore the need to develop improved models that achieve relatively low false-positive rates, enabling more routine testing and facilitating earlier disease detection with less risk of overdiagnosis.

Established risk calculators, such as the Prostate Cancer Prevention Trial (PCPT), combine PSA levels with other clinical factors and demographic information to estimate PCa risk, but do not currently incorporate PRSs. Recent studies have shown improved performance to these models by incorporating genetic information in the form of PRSs, which have shown promise in predicting both the risk of PCa incidence and the likelihood of mortality from the disease by stratifying men based on their genetic disposition of PCa. ^9^ Other studies have shown that the combined effect of other factors with PRSs such as carrier status for rare genetic variants, family history, and patient age at diagnosis have stronger associations with PCa incidence when compared to PRSs alone.^10^ However, the utility of PRSs can be influenced by factors such as the selection of single nucleotide polymorphisms (SNPs) used in their development, the diversity of the genome-wide association study (GWAS) cohort, the designated outcome of interest in the GWAS, and the generalizability to the applied cohort. While PRSs for PCa are associated with incident cancer and PCa mortality, recent research has shown that they do not enhance the prediction of aggressive cancers or outperform PSA testing.^11^ It is important to note that these studies do not take into account any social factors that could enhance the accuracy and clinical utility of PRS-based assessments for PCa. Therefore while the promise of PRSs is evident, its clinical application is still in the early stages, requiring further validation and integration with other risk factors to ensure its efficacy and utility.

### 2.2. Importance of Understanding Social Determinants for PCa Outcomes

While genetic differences can contribute to the development of certain diseases, there is a growing body of evidence suggesting that socioeconomic factors play a crucial role in disease outcomes. An analysis of temporal trends and key drivers of inequality in life expectancy revealed that behavioral and metabolic risk factors accounted for 74% of the county-level variation in life expectancy.^12^ Access to healthcare is a critical factor in accurately assessing disease outcomes, as demonstrated by two separate studies. Research by Moses et al. (2017) demonstrated that men from lower socioeconomic backgrounds are less likely to undergo regular PSA testing, leading to delayed diagnoses and poorer outcomes.^13^ In a Veterans Health Administration (VA) cohort study, Black men presented at a younger age with higher PSA levels. When treated in an equal-access setting, the disparity in PCa risk was significantly reduced, with Black men exhibiting an 11% lower risk of developing metastases compared to White men.^14^ Another study corroborated these findings by observing similar trends between Black and White men in the Surveillance, Epidemiology, and End Results (SEER) national cancer registry and the VA.^15^ These studies underscore the importance of addressing socioeconomic factors and ensuring equitable access to healthcare to reduce disparities in PCa outcomes and improve overall health equity.

## 3. Materials and Methods

### 3.1 Data Source

We used the All of Us (AoU) v7 Controlled tier dataset, a multi-domain database from the NIH’s Precision Medicine Initiative.^16^ The dataset integrates electronic health records (EHRs), physical measurements, genomics, and participants surveys. We analyzed three domains in the AoU dataset, the EHR, genomic, and survey domain.

### 3.2 Data Preparation

Participants were classified as cases if they had PCa, as indicated by the presence of relevant SNOMED or ICD9/ICD10 vocabulary codes. Additionally, these participants provided survey responses to the Basics, Lifestyle, Health Care Access & Utilization, Personal and Family Health History, and Social Determinants of Health surveys, as well as short-read whole genome sequencing data. Controls were identified as participants who had the same relevant data modalities but without a diagnosis of PCa. This comprehensive approach ensured that both cases and controls were well-characterized across multiple data sources. Beta coefficients for PCa susceptibility loci were obtained from the trans-ancestry GWAS conducted by Conti et al.^17^ These coefficients were utilized as the foundational weights for the construction of the PRSs.

### 3.2 PRS Development

Using the acquired beta coefficients, we developed polygenic risk scores (PRS) for approximately 2,556 PCa cases and 21,974 controls. This process involved calculating individual-level risk scores based on the sum of risk allele counts weighted by their respective beta coefficients. Weight files were converted to the GRCh38 reference assembly using LiftOver to ensure compatibility with the reference genome used in the AoU genomic dataset. Quality control checks such as for heritability, effect allele designation, and relatedness of the data were performed to remove unwanted samples. The ORs from 269 risk variants identified in the Conti et al study were used to calculate the raw PRSs. Raw PRSs were adjusted using residualization prior to being used in the modeling of PCa incidence.

### 3.3 Data Integration and Processing

Following the development of the PRSs, these scores were integrated with a comprehensive array of survey and health data to enhance predictive accuracy. We utilized surveys from the AoU, including The Basics, Overall Health, Lifestyle, Personal and Family Health History, and Health Care Access and Utilization. In total, 11 questions from these surveys were employed to inform the relevant categories (Table 1). Additionally, age, zip code, and socioeconomic variables were derived from the relevant tables provided by AoU. Body mass index (BMI) was calculated for all participants based on the physical measurements data collected through AoU and included as a biometric in the analysis. Furthermore, the binary presence of comorbidities—such as hypertension, diabetes, stroke, and high cholesterol—was extracted using ICD9/10 source concept codes. The final cohort included 2,317 White cases with 19,056 controls, 189 Black or African American cases with 1,976 controls, and 23 Asian cases with 586 controls. Despite low counts in some categories, participants from Middle Eastern or North African (116), Multiracial (267), Hispanic White (241), Hispanic Black (26), and Hispanic Other (33) groups were included to enhance representativeness.

**Table 1.** Importance of Features by Survey Type.

| Survey | Questions | Model Feature Importance |
| --- | --- | --- |
| Basics | • What is the highest grade or year of school you completed?<br>• Are you covered by health insurance or some other kind of health care plan?<br>• What is your current employment status? Please select 1 or more of these categories.<br>• What is your annual household income from all sources?<br>• In the past 6 months, have you been worried or concerned about NOT having a place to live? | Medium |
| Family Health History | • Who in your family has had prostate cancer? Select all that apply. | High |
| Health Care Access and Utilization | • Is there a place that you USUALLY go to when you are sick or need advice about your health?<br>• About how long has it been since you last saw or talked to a doctor or other health care provider about your own health? | Medium |
| Lifestyle | • Have you smoked at least 100 cigarettes in your entire life? | Low |
| Social Determinants of Health | • Within the past 12 months, we worried whether our food would run out before we got money to buy more. | Low |

### 3.3 Model Training and Evaluation

To address the substantial class imbalance between cases and controls, we applied the Synthetic Minority Over-sampling Technique (SMOTE) from the imblearn library to the training set. SMOTE generates synthetic samples to enhance the representation of the minority class, thereby improving model training outcomes. We used cross-validation with 5 folds to assess the model’s performance consistently. The dataset was divided into an 80/20 ratio for training and test sets for each fold. This split was crucial for developing and validating the logistic regression (LR) model used to predict PCa risk. We employed a logistic regression model to estimate the odds ratios (ORs) for each feature. The average false positive rate (FPR) across all folds was computed to provide a robust measure of the model’s performance. The importance of each feature in the model was analyzed to understand its contribution to PCa risk prediction (Figure 2).

**Figure 1.**
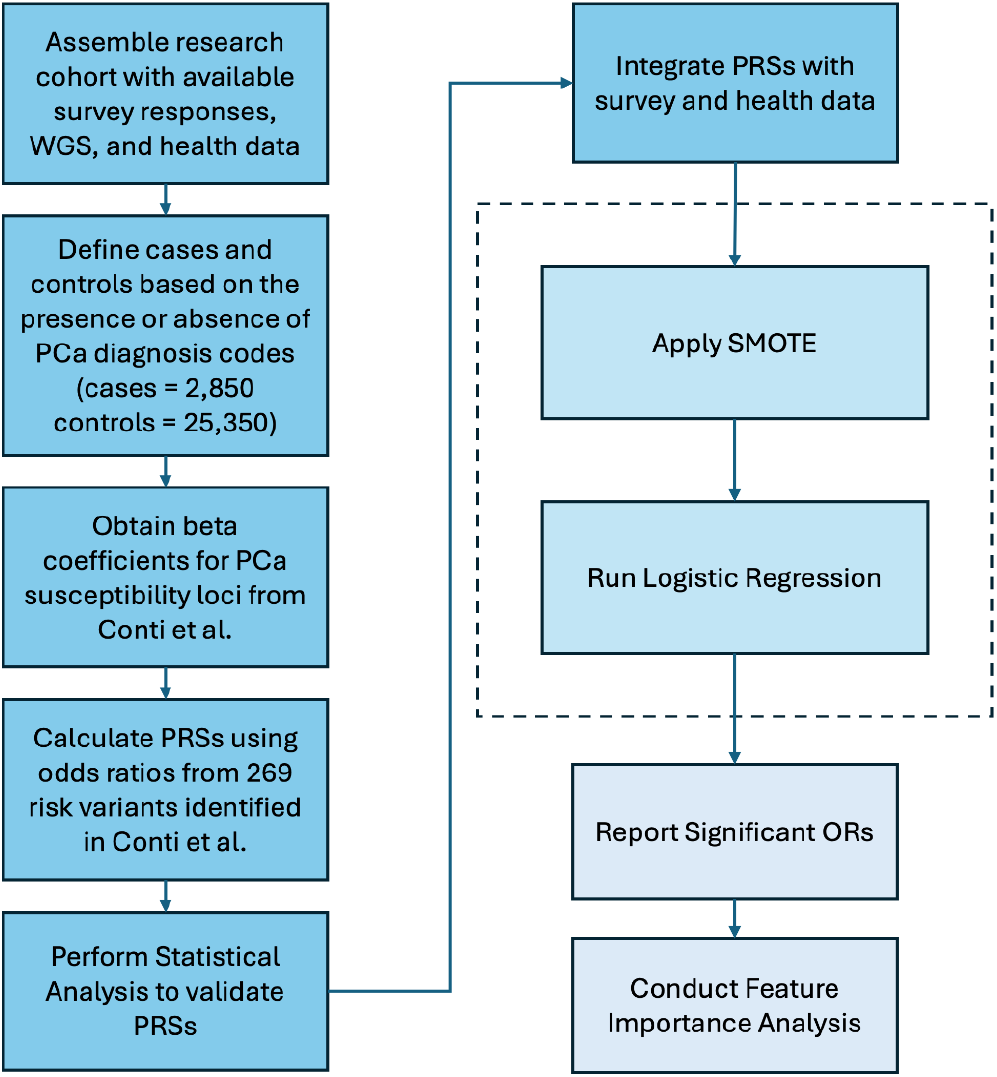
Data Processing and Model Workflow

**Figure 2.**
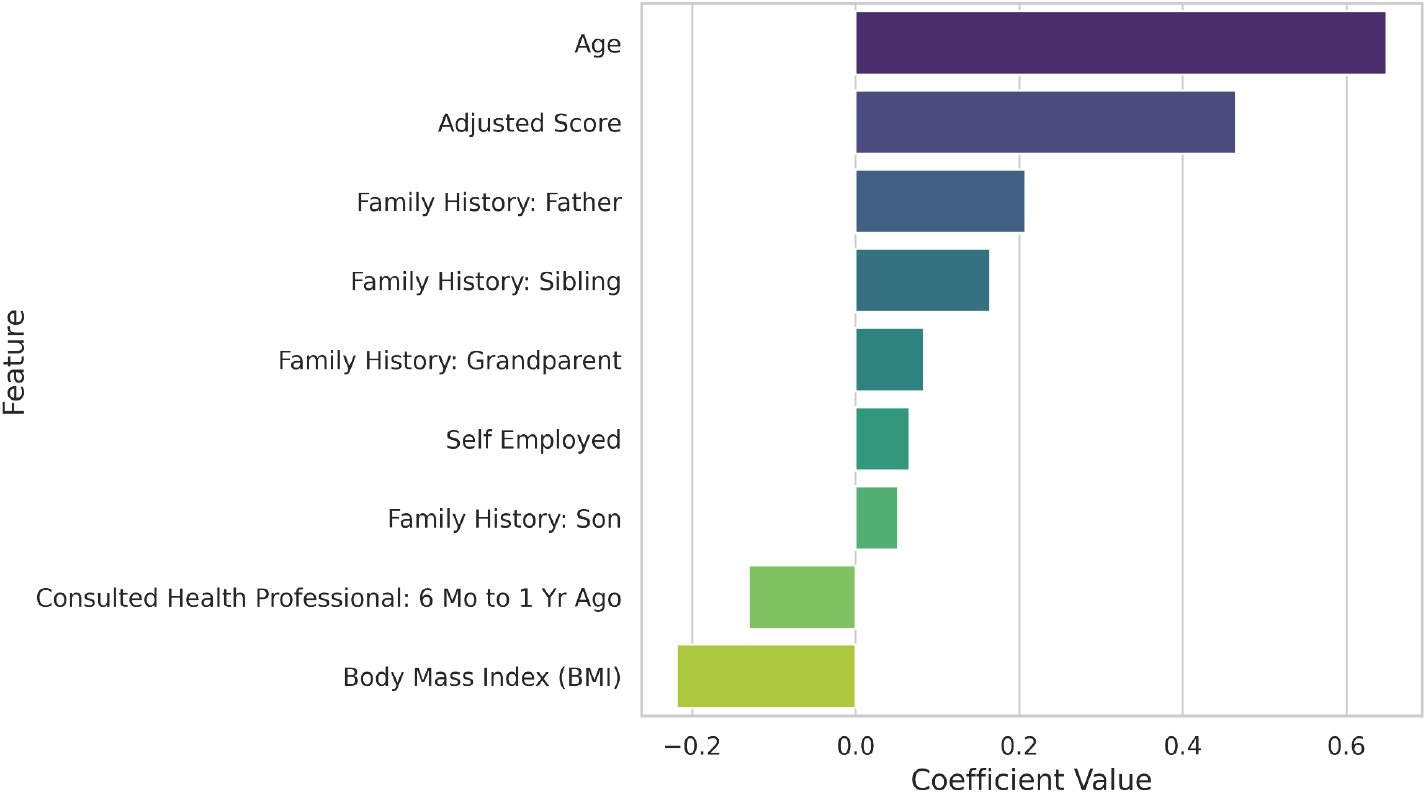
Feature Importance of Significant Variables from Logistic Regression

**Figure 3.**
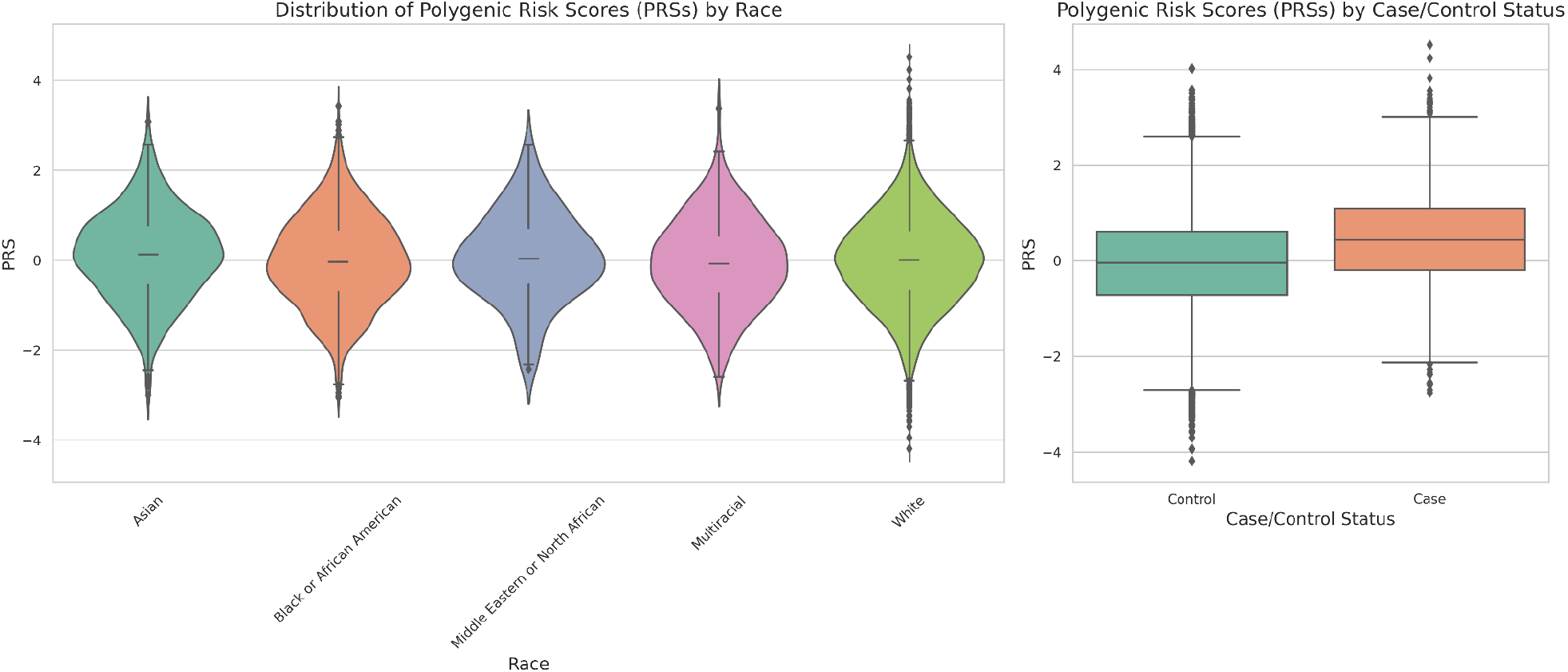
Distribution of Polygenic Risk Scores by different subgroups

## 4. Results

### 4.1 Full Multimodal Model

In the analysis of PCa incidence, several variables demonstrated significant associations with the outcome. A family history of PCa in a father (OR = 1.23, p < 0.001), grandparent (OR = 1.23, p < 0.001), sibling (OR = 1.18, p < 0.001), and son (OR = 1.05, p = 0.001) was strongly linked to increased risk. Age also emerged as a significant predictor, with each additional year increasing the odds of PCa by 1.92 (p < 0.001). A higher BMI was associated with a decreased risk (OR = 0.80, p < 0.001). The adjusted PRS score positively influenced the odds of PCa (OR = 1.59, p < 0.001). Additionally, being self-employed was associated with a marginal increase in risk (OR = 1.07, p = 0.032), while having consulted a health professional 6 months to 1 year ago was linked to a decreased risk (OR = 0.88, p = 0.018). These findings highlight the interplay between genetic, socio-economic, and health-related factors in influencing PCa risk.

After applying SMOTE for resampling, the logistic regression model achieved an area of the receiver operating characteristic curve (AUROC) of 0.79. It correctly identified 403 PCa cases, though 139 cases were misclassified. The model also accurately classified 3,428 non-cancer cases, with 1,406 false positives which represents a 29% FPR. Precision for cancer cases was 0.22, and recall was 0.74, indicating a strong ability to detect cancer cases but with room for improvement in precision.

### 4.2 Model Performance Across Different Feature Subsets

The performance of models incorporating different feature subsets varied across key metrics. The model using only age and polygenic risk scores (PRS) achieved an AUROC of 0.75, with a recall of 0.72, precision of 0.19, and did not predict any positive cases. Introducing health metrics to the feature set improved AUROC to 0.80 and recall to 0.74, though precision slightly increased to 0.22, and the FPR rose to 0.29. Including race data alongside age, PRS, and health metrics resulted in a marginal decrease in AUROC to 0.79 and recall to 0.69, while precision increased to 0.23, and the FPR decreased to 0.26. Although non-cancer classifications were robust, all models faced significant challenges in accurately detecting PCa, underscoring the need for further refinement to enhance sensitivity and reduce false positives.

Statistical analysis of PRSs between cases and controls revealed a significant difference. The independent t-test and Mann-Whitney U test both yielded p-values less than 0.001. This indicates a substantial disparity in PRSs between the PRS distribution for PCa cases and controls. An Analysis of Variance (ANOVA) was conducted to evaluate differences in PRSs across different racial groups. The results indicated that there was no statistically significant difference in PRS scores among the racial groups.

## 5. Discussion

In this study, we investigated the complexity of PCa screening using the national All of Us research cohort. We identified several key factors influencing PCa risk, including increased age, family history, and higher PRSs. Additionally, we found that recent healthcare consultations were associated with a reduced risk of PCa, potentially highlighting the role of healthcare access and early detection. The adjusted PRS emerged as a significant predictor of PCa, supporting the utility of genetic information in screening strategies. However, the inclusion of SDoHs, such as employment status and recent healthcare consultations, also demonstrated notable associations with PCa risk. These findings underscore the importance of considering genetic, family health history, and healthcare utilization factors in PCa risk assessment. By advancing our understanding of these diverse factors, this research contributes to overcoming health disparities in precision medicine, emphasizing the need for tailored strategies to improve cancer prevention and treatment outcomes across different populations.

Notably, we discovered that individuals who visited a healthcare professional within the past 6 months to a year exhibited a reduced risk of PCa compared to those who consulted a professional at other frequencies. This highlights important factors associated with PCa that differ from other major studies which primarily focus on genetic and biological factors.^18^ Regular consultations likely facilitate early detection and intervention and may be related to insurance benefits, such as improved insurance coverage and affordability which could influence healthcare utilization and early detection. Addressing barriers to healthcare access can mitigate disparities in PCa outcomes, emphasizing the need for equitable healthcare services. These findings highlight the importance of healthcare utilization in cancer prevention and the broader implications for health equity.

Our study identified age, higher BMI (associated with decreased risk), employment status (being self-employed), and recent healthcare consultations as significant predictors of PCa risk. These findings align with other major studies, reinforcing the reliability and reproducibility of these predictors in understanding PCa risk.^19^ Particularly, the association between employment status and healthcare access aligns with existing literature, highlighting the multifaceted nature of PCa risk factors.^20^ The associations between self-employment and marginally increased risk of PCa incidence may potentially reflect variations in occupational stress, health behaviors, or healthcare access among different employment types. Additionally, the seemingly paradoxical association between higher BMI and a decreased risk of PCa may be attributed to detection bias. Obese men often have lower PSA levels, which is believed to result from hemodilution due to their larger blood volume and increased prostate size. This dilution reduces the concentration of PSA in the bloodstream, making it more challenging to accurately detect existing cancer.^21^ While all of the mechanistic players in this relationship are not yet fully understood, they can potentially involve complex biological pathways, which could influence PCa progression differently compared to other cancers.

Adjustment of class instances in a model can have a strong influence on model performance and outcomes. Ideally, with a sufficiently large dataset, the reliance on data generation techniques like SMOTE could be minimized, alleviating concerns about models trained on synthetic data. However, developing a robust framework for testing with real-world data remains essential. In our case, using SMOTE significantly improved model recall, but it also led to a higher false positive rate (FPR). Further research is needed to explore alternative methods for addressing class imbalance and their impact on FPR.

Our models were robust. We evaluated several models using different combinations of feature subsets, and found that the full multimodal model incorporating PRSs, health data, and surveys outperformed the others. Our analysis revealed a progressive enhancement in model performance with the inclusion of data from diverse modalities. Models without social data showed limited accuracy in predicting the positive class. With the race variable included in the full model, we observed challenges with recall. Although the model with race included achieves a slightly higher FPR, we know that race is confounded by multiple SDoH variables which does not clearly allow for assessment of race’s importance in the model. The drop in recall performance for the model with race suggests that the inclusion of race may have led the model to overly prioritize this factor, potentially overshadowing other critical features and contributing to increased misclassification rates. Future work includes looking at the interaction of race with a wide range of social data available. These findings underscore the importance of adopting a comprehensive approach that considers multiple variables beyond race to enhance both the robustness and fairness of predictive models.

## 6. Limitations

The analysis was limited by the lack of PSA data and Gleason scores, crucial biomarkers for PCa diagnosis and management. To address this limitation, we relied on other available risk factors, such as polygenic risk scores and socio-economic determinants, to build our predictive model. Additionally, the study could not evaluate the model’s ability to predict PCa mortality due to insufficient mortality data. We addressed this limitation by focusing on incidence rather than mortality, but this approach limits the scope of our findings. Future research should incorporate mortality data to provide a more comprehensive assessment of PCa prognosis.

Some surveys (The Basics, Lifestyle, and Overall Health) are mandatory and completed at baseline, defined as the time of registration, and regularly thereafter. The remaining surveys are available as follow-ups. Baseline surveys, which capture information such as employment status, can thus be used to assess a participant’s status prior to a PCa diagnosis, as only diagnoses occurring after registration were included in the analysis. However, follow-up surveys have a substantially lower response rate compared to baseline surveys and may not accurately reflect a participant’s status before a PCa diagnosis. To address this, only surveys strictly pertaining to the period before diagnosis could be included in the analysis, though this approach would severely reduce the number of individuals included, particularly among underrepresented racial groups. Further research is needed to address this limitation.

## 7. Conclusions

This study highlights the complex interplay between genetic factors, socio-economic determinants, and health behaviors in influencing PCa risk and introduces a novel approach by integrating data from the three respective domains in the AoU research program. Unlike traditional risk calculators for PCa, we predicted incident PCa without utilizing race or PSA data and achieved performance comparable to traditional models. While family history and age emerged as significant predictors, the influence of PRS and socio-economic factors underscores the need for a multimodal screening strategy. Despite achieving moderate accuracy, the models’ high false positive rate and variable performance across feature subsets indicate the necessity for further refinement. These findings advocate for a balanced approach in PCa screening that incorporates both genetic and socio-economic dimensions, aiming to enhance precision, reduce overdiagnosis, and promote health equity. This work establishes a methodological framework for incorporating SDoH data into clinical risk assessments for PCa and potentially other cancer types, enhancing the comprehensiveness and equity of predictive modeling in clinical settings.

## Data Availability

All data produced in the present study are available upon reasonable request to the authors

https://github.com/ashlew23/pc_multimodal

## 8. Acknowledgements

This work was supported in part by the Blavatnik Fellowship. We thank the Department of Biomedical Data Science for resources to support this effort. We gratefully acknowledge All of Us participants for their contributions, without whom this research would not have been possible. We also thank the National Institutes of Health’s All of Us Research Program for making available the participant data examined in this study.

## Footnotes

1 Mateo et al., “Accelerating Precision Medicine in Metastatic Prostate Cancer.”

2 Loeb et al., “Overdiagnosis and Overtreatment of Prostate Cancer.”

3 Cuzick et al., “Prevention and Early Detection of Prostate Cancer.”

4 Black et al., “Validation of a Prostate Cancer Polygenic Risk Score.”

5 Schaffer et al., “A Polygenic Risk Score for Prostate Cancer Risk Prediction.”

6 Mohsen et al., “Artificial Intelligence-Based Methods for Fusion of Electronic Health Records and Imaging Data.”

7 US Preventive Services Task Force, “Screening for Prostate Cancer.”

8 Tosteson et al., “Consequences of False-Positive Screening Mammograms”; Lin et al., “Benefits and Harms of Prostate-Specific Antigen Screening for Prostate Cancer.”

9 Klein et al., “Prostate Cancer Polygenic Risk Score and Prediction of Lethal Prostate Cancer.”

10 Darst et al., “Combined Effect of a Polygenic Risk Score and Rare Genetic Variants on Prostate Cancer Risk”; Zheng et al., “Genetic Variants and Family History Predict Prostate Cancer Similar to Prostate-Specific Antigen”; Na et al., “Single-Nucleotide Polymorphism-Based Genetic Risk Score and Patient Age at Prostate Cancer Diagnosis.”

11 Schaffer et al., “A Polygenic Risk Score for Prostate Cancer Risk Prediction”; Klein et al., “Prostate Cancer Polygenic Risk Score and Prediction of Lethal Prostate Cancer.”

12 Dwyer-Lindgren et al., “Inequalities in Life Expectancy Among US Counties, 1980 to 2014.”

13 Moses et al., “The Impact of Sociodemographic Factors and PSA Screening Among Low Income Black and White Men.”

14 Yamoah et al., “Racial and Ethnic Disparities in Prostate Cancer Outcomes in the Veterans Affairs Health Care System.”

15 Klebaner, Courtney, and Rose, “Effect of Healthcare System on Prostate Cancer-Specific Mortality in African American and Non-Hispanic White Men.”

16 “Data Sources – All of Us Research Hub.”

17 Conti et al., “Trans-Ancestry Genome-Wide Association Meta-Analysis of Prostate Cancer Identifies New Susceptibility Loci and Informs Genetic Risk Prediction.”

18 Schaffer et al., “A Polygenic Risk Score for Prostate Cancer Risk Prediction”; Black et al., “Validation of a Prostate Cancer Polygenic Risk Score”; Klein et al., “Prostate Cancer Polygenic Risk Score and Prediction of Lethal Prostate Cancer.”

19 Buschemeyer and Freedland, “Obesity and Prostate Cancer”; Porter and Stanford, “Obesity and the Risk of Prostate Cancer”; Penson et al., “The Association between Socioeconomic Status, Health Insurance Coverage, and Quality of Life in Men with Prostate Cancer”; Mahal et al., “The Association between Insurance Status and Prostate Cancer Outcomes.”

20 Lee et al., “Unmet Healthcare Needs Depending on Employment Status”; McMaughan, Oloruntoba, and Smith, “Socioeconomic Status and Access to Healthcare.”

21 Buschemeyer and Freedland, “Obesity and Prostate Cancer.”

